# Umbilical Cord Blood DNA Methylation in Children Who Later Develop Type 1 Diabetes

**DOI:** 10.1101/2021.05.21.21257593

**Authors:** Essi Laajala, Ubaid Ullah, Toni Grönroos, Omid Rasool, Viivi Halla-aho, Mikko Konki, Roosa Kattelus, Juha Mykkänen, Mirja Nurmio, Mari Vähä-Mäkilä, Henna Kallionpää, Niina Lietzén, Bishwa R. Ghimire, Asta Laiho, Heikki Hyöty, Laura L. Elo, Jorma Ilonen, Mikael Knip, Riikka J. Lund, Matej Orešič, Riitta Veijola, Harri Lähdesmäki, Jorma Toppari, Riitta Lahesmaa

**Author notes:** equal contribution (Jorma Ilonen, Mikael Knip, Riikka J. Lund, Matej Orešič, Riitta Veijola). **Correspondence** Professor Riitta Lahesmaa, Turku Bioscience Centre, Tykistökatu 6A, Turku 20520, FINLAND.

## Abstract

Distinct DNA methylation patterns have recently been observed to precede type 1 diabetes in whole blood collected from young children. Our aim was to determine, whether perinatal DNA methylation could be associated with later progression to type 1 diabetes. Reduced representation bisulfite sequencing (RRBS) analysis was performed on umbilical cord blood samples collected within the Type 1 Diabetes Prediction and Prevention (DIPP) study. Children later diagnosed with type 1 diabetes and/or testing positive for multiple islet autoantibodies (N=43) were compared to control individuals (N=79), who remained autoantibody-negative throughout the DIPP follow-up until 15 years of age. Potential confounding factors related to the pregnancy and the mother were included in the analysis. No differences in the cord blood methylation patterns were observed between these cases and controls.

## INTRODUCTION

DNA methylation at cytosine residues is one of the most important epigenetic mechanisms regulating gene expression. The modification converts cytosine to 5-methylcytosine, usually in the context of CpG dinucleotides. Differential methylation at the promoter or other regulatory elements affect gene expression in health and diseases, e.g. type 2 diabetes (1) and systemic lupus erythematosus (2). Most studies on the association between type 1 diabetes and DNA methylation have focused on differences between case and control subjects at the time of diagnosis or later. For example, methylation patterns at the promoters of insulin and IL2RA genes have been associated with type 1 diabetes (3,4). The most extensive study on the topic included immune effector cells from 52 monozygotic twin pairs discordant for type 1 diabetes (5). Thousands of CpG-sites were found to be differentially variable between affected subjects and their healthy co-twins. An earlier study by the same group (6) included a small set of samples from pre-diabetic individuals (N=7) to confirm their findings from already-diagnosed subjects. A recent report from the DAISY study on prospective epigenomics of type 1 diabetes also included samples collected before the case subjects’ seroconversion to islet autoantibody positivity (7). They discovered differential methylation at 30 genomic regions between longitudinal samples of 87 autoantibody-negative control individuals and 87 case individuals who later progressed to type 1 diabetes.

DNA methylation patterns are largely established *in utero* (8), and neonatal DNA methylation marks have been associated with later health outcomes (9,10). We therefore hypothesized, that the progression to type 1 diabetes during childhood might be reflected in the epigenome already at birth. Two earlier studies have examined umbilical cord blood samples from neonates, who later progressed to type 1 diabetes (5,7). In both studies, the neonatal samples were only utilized to confirm the direction of change in the differentially methylated regions discovered at later time points. However, they did not publish umbilical cord blood DNA methylation measurements outside the candidate regions, and neither were the studies designed to compare neonatal samples.

In this study we used the reduced representation bisulfite sequencing (RRBS) method to analyze umbilical cord blood DNA methylation associated with later progression to type 1 diabetes in a prospective cohort, the Finnish Type 1 Diabetes Prediction and Prevention (DIPP) study. The aim was to detect DNA methylation patterns associated with later progression to type 1 diabetes. Such findings would be valuable for a better understanding of early mechanisms underlying the progression to type 1 diabetes related autoimmunity.

## RESEARCH DESIGN AND METHODS

The supplementary material includes a more detailed description of the sample collection protocol, HLA risk class determination (11), the bisulfite sequencing and the statistical methods.

### Study design

Case children (N = 43) who were diagnosed with type 1 diabetes during the DIPP follow-up or became persistently positive for at least two biochemical islet autoantibodies (in at least two consecutive serum samples) were compared to control children (N = 79) who remained autoantibody-negative throughout the DIPP follow-up, i.e. up to 15 years of age or until their decision to discontinue participation in the DIPP study. Data until the end of year 2018 were included. Clinical data such as maternal insulin-treated diabetes, gestational weight gain, and the child’s birth weight were utilized to adjust for potential confounding effects. The characteristics of the case and control children are described in Table 1.

**Table 1:**
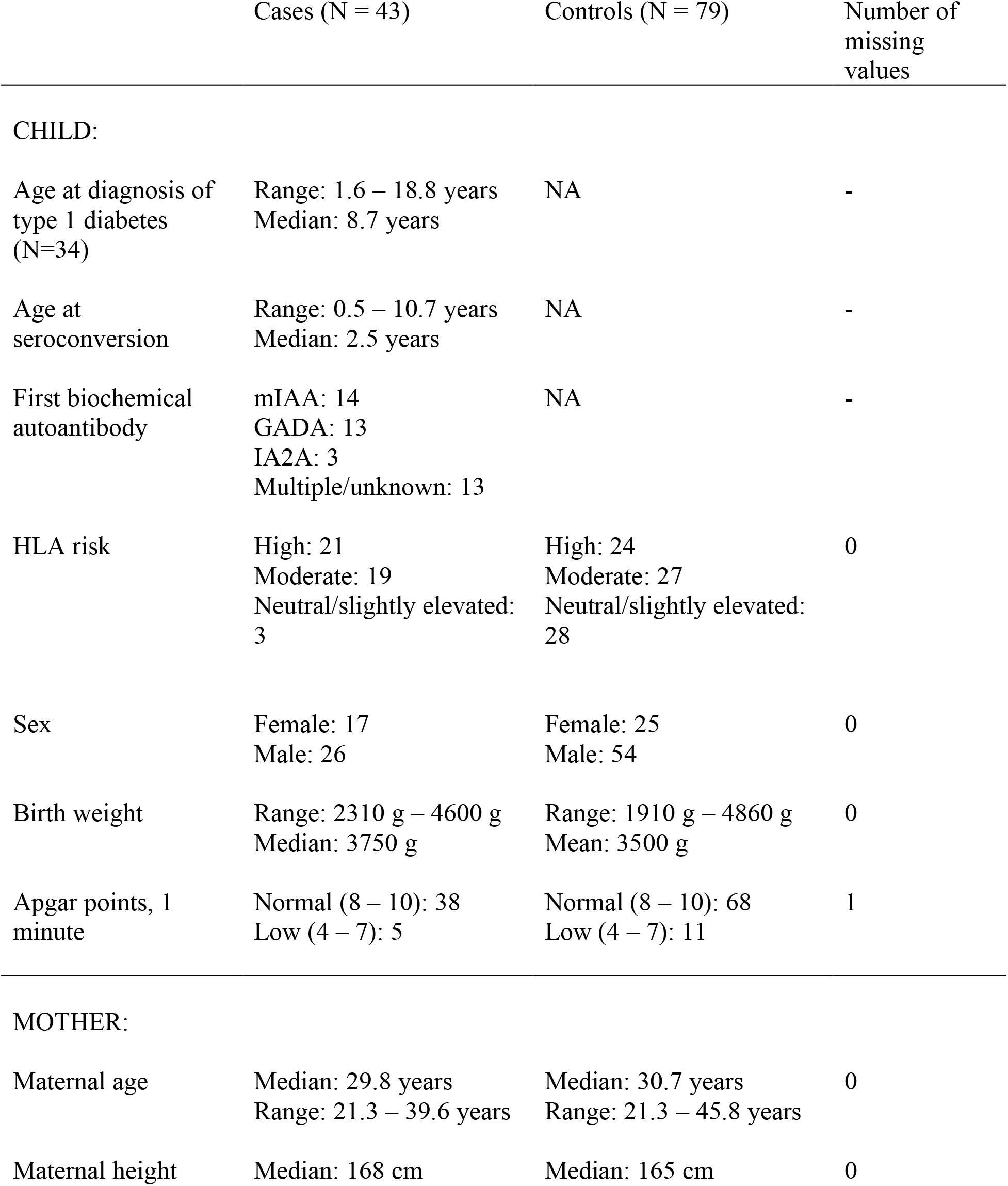
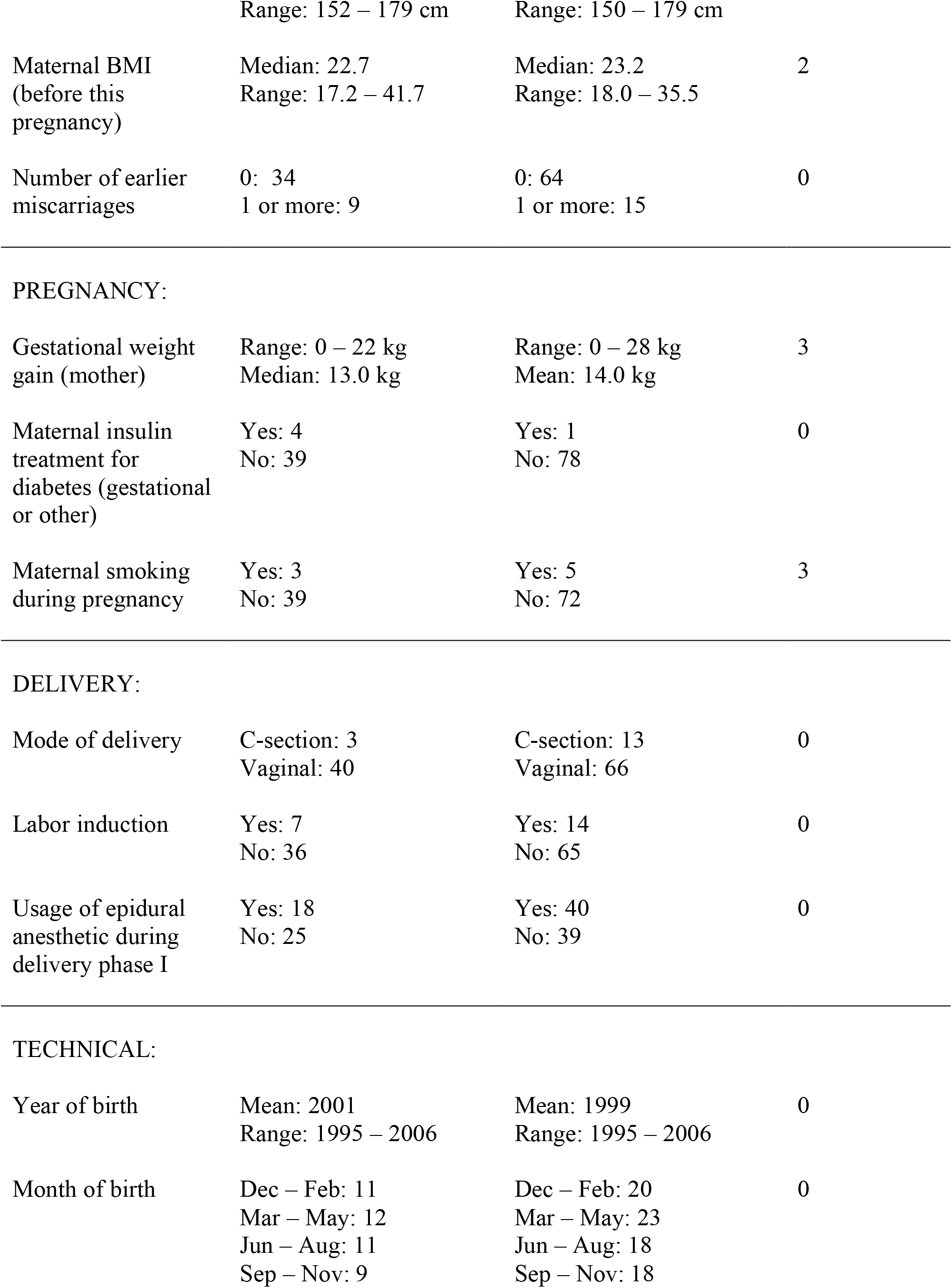
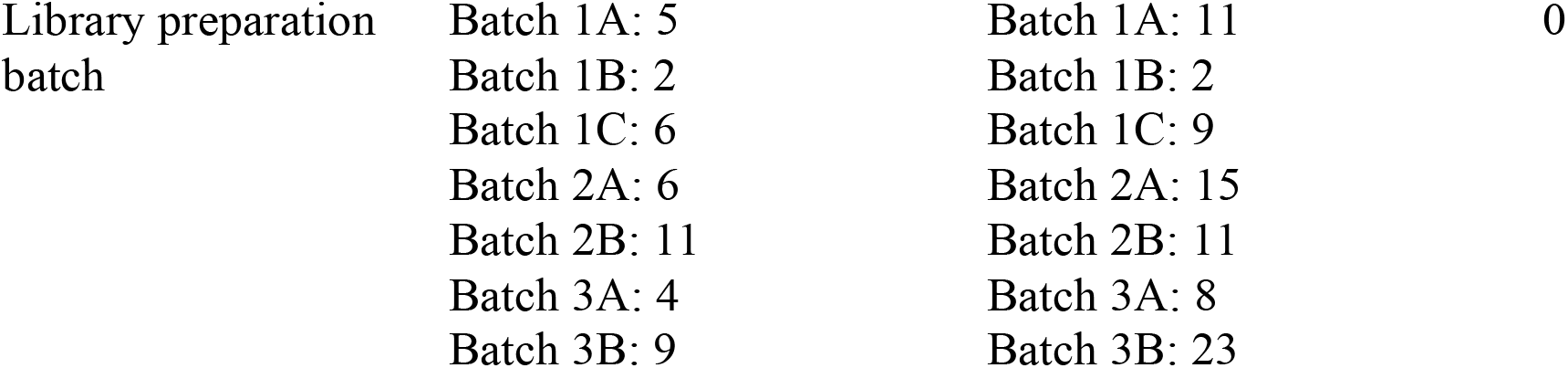
Characteristics of the case and control children. All the covariates listed here were included as explanatory variables in the differential methylation analysis, except for age at diagnosis, age at seroconversion and first-appearing autoantibody, which are relevant only for the case group. The inclusion criteria are specified in the Supplementary material.

Umbilical cord blood samples were collected from newborn children born in Turku University Hospital between 1995 and 2006. After informed consent, HLA DR/DQ genotyping was performed from umbilical cord blood to identify children at increased risk to develop type 1 diabetes. Eligible children were invited to participate in the DIPP follow-up. Criteria for eligibility have been modified during the study years to increase the sensitivity and specificity of the screening (11). During DIPP follow-up, serum samples for islet autoantibody measurements were collected at 3-month intervals until the age of 2 years and thereafter every 6 or 12 months until the age of 15 years or until type 1 diabetes diagnosis. Islet autoantibodies were measured with specific radio-binding assays and included IAA (insulin autoantibody), IA-2A (insulinoma-associated protein 2 antibody), GADA (glutamic acid decarboxylase antibody) and ZnT8A (zinc transporter-8 antibody). Classical islet cell antibodies (ICA) were used as the only autoantibody screening method for DIPP children born until the end 2002, and if positive, all other autoantibodies were measured from all earlier and future samples of the child. ICA, IAA, GADA and IA-2A were measured from all follow-up samples from children born since 2003, whereas ZnT8A were measured only if at least one of the other autoantibodies became positive (12). In addition, all five islet autoantibodies were measured from all samples of the first 1000 children participating in DIPP follow-up (12).

### DNA methylation profiling

The library preparations steps were adapted from the RRBS protocol by Boyle et al. (13). Illumina HiSeq 2500 instrument was used for paired-end sequencing (2 x 100 bp) of the DNA libraries. TrimGalore (14) and Bismark (15) were used for trimming and alignment of paired-end RRBS reads on the GRCh37 (hg19) genome assembly (16). After removing M-biases, potential single nucleotide polymorphisms (17), and sites with extremely high coverages, a minimum coverage of 10 reads was required for at least one third of the samples in both groups.

### Differential methylation analysis

A generalized mixed effects model (GLMM) implemented in the R package PQLseq (18) was fit separately for read counts at each CpG site on autosomal chromosomes. The covariates listed in Table 1 were modeled as fixed effects and the genetic similarity between individuals as a random effect. Since we could not know, which covariates might be important confounding factors, we included all available reliably recorded information. However, only one covariate was selected from each group of mutually correlated clinical covariates (detailed inclusion criteria are listed in Supplementary Table 1). A few missing covariate values were median-imputed, and continuous covariates were Z-transformed. The Wald test P values computed within PQLseq were spatially adjusted by a weighted Z-test implemented in package RADMeth (19). Since the spatially adjusted P values were found to be inflated, false discovery rate (FDR) was estimated empirically through a permutation analysis (20).

### Pyrosequencing validation of selected targets

Technical validation was performed by targeted pyrosequencing with PyroMark Q24 system (Qiagen) using 58 samples, which were a subset of the samples studied with RRBS. We chose an even number of male and female case and control subjects, who were born vaginally at full term with normal birth weight and normal Apgar score and were not exposed to maternal smoking in utero. The genomic regions of interest were captured with an assay and amplified by 45 rounds of PCR. An ordinary linear regression model was fit for each DNA methylation proportion, after applying a transformation: arcsin(2 × proportion – 1). The included explanatory variables are specified in the supplementary material. Each model was fit with and without the covariate of interest, and the significance of the association was estimated based on an ANOVA test comparing the two models.

### Ethical aspects

All participating families gave an informed consent for the genetic HLA screening from umbilical cord blood and for the follow-up. The study was originally approved by the Ethics Committee of the Hospital District of Southwest Finland followed by the Ethics Committee of the Hospital District of Northern Ostrobothnia. The study followed the principals of the Helsinki II declaration.

### Data and resource availability

The datasets generated during the current study will be available in a public repository as soon as the manuscript is accepted for publication. The code is available in https://github.com/EssiLaajala/RRBS_workflow, where also the data availability status will be updated.

## RESULTS

Altogether 2568146 CpG sites fulfilled the quality and coverage criteria and were included in the differential methylation analysis. None of them were differentially methylated between the case and the control children as individual CpG sites (Benjamini-Hochberg-corrected Wald test P value < 0.05 before the spatial adjustment). After spatial adjustment, two adjacent CpG sites (chr11:400288 and chr11:400295, GRCh37 genome assembly) on an intron of gene Plakophilin-3 (*PKP3*) showed weak evidence of hypomethylation in the case children, as compared to the controls (Table 2). However, technical replication by targeted pyrosequencing showed that the difference was not significant (Figure1A-B, Supplementary Figure 1).

**Table 2:**
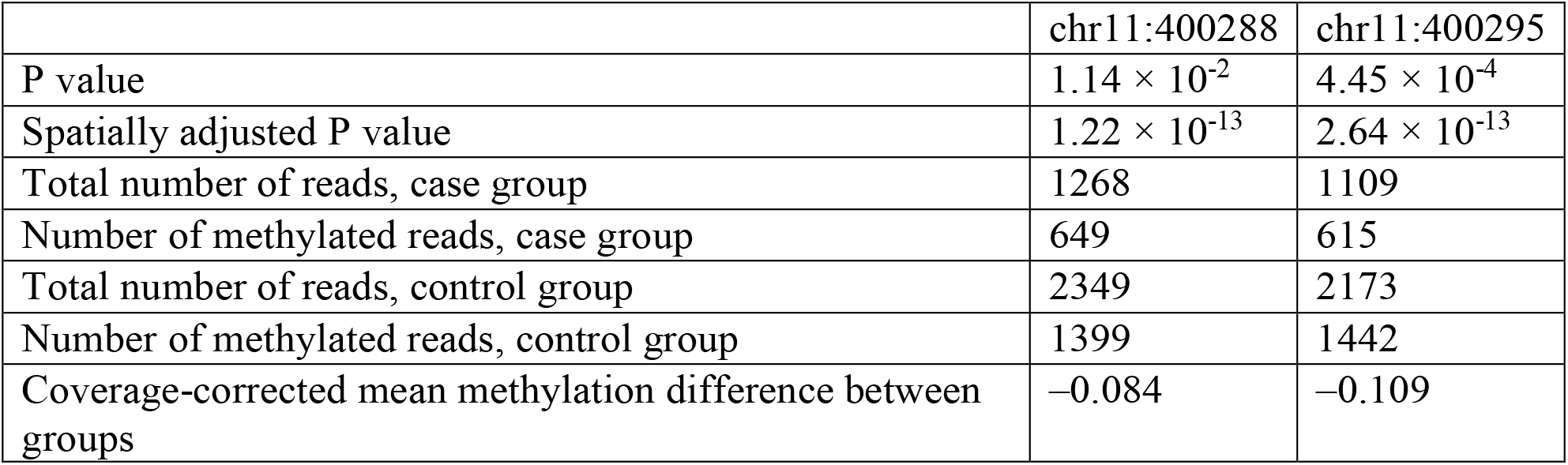
Two adjacent CpG sites on the intron of *PKP3* (GRCh37 genome assembly) showed weak evidence of hypomethylation in the case group (N=43), as compared to the control group (N=79). Coverage-corrected mean methylation difference is calculated as sum(number of methylated reads in cases) / sum(number of total reads in cases) – sum(number of methylated reads in controls) / sum(number of total reads in controls).

|  | chr11:400288 | chr11:400295 |
| --- | --- | --- |
| P value | $1.14 \times 10^{-2}$ | $4.45 \times 10^{-4}$ |
| Spatially adjusted P value | $1.22 \times 10^{-13}$ | $2.64 \times 10^{-13}$ |
| Total number of reads, case group | 1268 | 1109 |
| Number of methylated reads, case group | 649 | 615 |
| Total number of reads, control group | 2349 | 2173 |
| Number of methylated reads, control group | 1399 | 1442 |
| Coverage-corrected mean methylation difference between groups | -0.084 | -0.109 |

**Figure 1:**
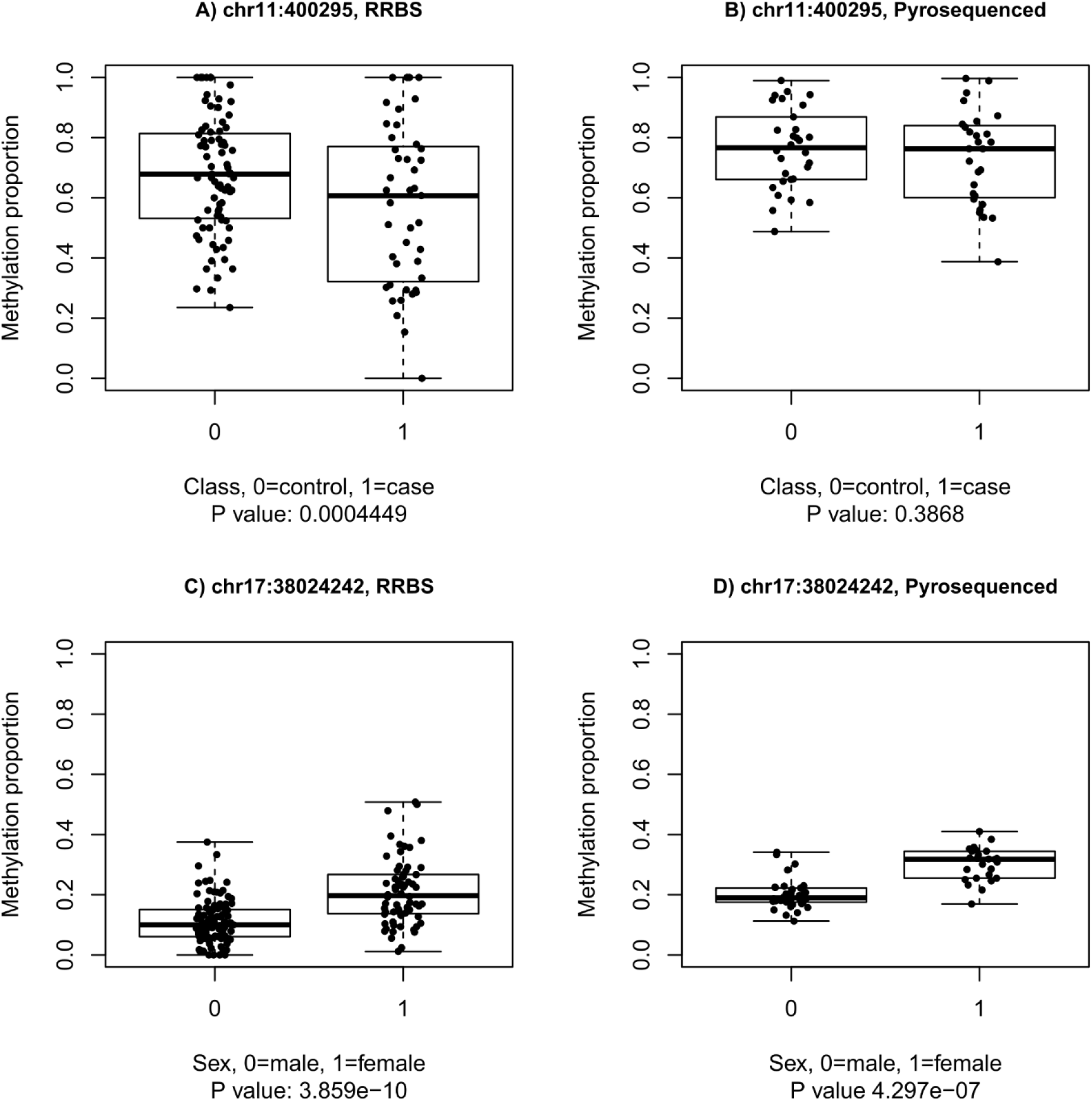
Methylation proportions quantified with two different technologies (RRBS and targeted pyrosequencing) at two example CpG sites visualized as boxplots (similar figures for all pyrosequenced targets are presented in the Supplementary material). A CpG site at Chr11:400295 (A-B) on an intron of Plakophilin 3 (*PKP3*) showed weak evidence of differential methylation between case and control subjects (not as an individual cytosine but as part of a candidate differentially methylated region), as measured by RRBS (A) and pyrosequencing (B). A CpG site at Chr17:38024242 (C-D) on the promoter of Zona Pellucida Binding Protein 2 (*ZPBP2*) was differentially methylated (genome-wide significance) between the sexes as an individual CpG site and as part of a differentially methylated region, as quantified by RRBS (C). Targeted pyrosequencing confirmed the differential methylation at this location (D), as well as at other CpG sites in the region (Supplementary Material). The P values marked below each plot are nominal (neither spatially adjusted nor multiple testing corrected). The midline of each boxplot is drawn at the median, boxes range from the 1^st^ to the 3^rd^ quartile, and whiskers extend to the most extreme values.

The strong inflation of spatially adjusted P values was an important observation in these data, as described by Laajala et al. (20). Before the inflation was discovered, 28 genomic regions were considered differentially methylated between cases and controls, based on Benjamini-Hochberg-corrected spatially adjusted P values (< 0.05). We carried out pyrosequencing to validate five selected targets technically, but the results did not indicate differential methylation between the groups (Supplementary Table 2). Empirical false discovery rate control of the RRBS results further confirmed that the differences were indeed not significant.

We selected a sex-associated differentially methylated region as a positive control to confirm that concordant results could be obtained by two different technologies (RRBS and targeted pyrosequencing) in the analyzed samples. This target (chr17:38024242–38024291) included six CpG sites that were hypomethylated in males (N=105), as compared to females (N=68). The region is located on the promoter of Zona pellucida binding protein 2 (*ZPBP2*), whose expression is highly specific to the testis tissue (GTEx Portal on 03/02/21). Pyrosequencing results confirmed differential methylation between sexes with P values in the order of 10^−6^–10^−9^ at each CpG site (Supplementery Figure 2). One of these CpG sites is visualized in Figure1C-D.

## DISCUSSION

Distinct DNA methylation patterns have recently been observed to precede type 1 diabetes in whole blood collected from very young children (7). We tested the possible presence of such differences already at the time of birth in a collection of umbilical cord blood samples. Compared to earlier studies, our data covered a substantially larger number of CpG sites and was specifically designed to compare DNA methylation at the time of birth. Based on our results, differences between children who progress to type 1 diabetes and those who remain healthy throughout childhood, are not yet present in the perinatal DNA methylome.

As several earlier studies have already pointed out, effect sizes in umbilical cord blood DNA methylation studies are small (21,22). The median methylation difference between males and females at sex-associated differentially methylated cytosines (DMCs) was 10 %, range 0.6 % - 35.2 % (20). The DMCs selected for technical validation with pyrosequencing, all of which were successfully validated, had a median methylation difference of 11.6 % between the sexes in the RRBS data. Since the sample numbers were relatively large in all comparisons (we compared 68 female individuals to 105 male individuals and 43 case individuals to 79 control individuals), effects of the class covariate of similar sizes probably would have been detected if they had been present in these data. However, we cannot exclude the possibility that such differences could be found in a larger data set.

A peak in the appearance of islet autoimmunity occurs at an early age, between one and two years (23,24), suggesting an early trigger that could act already before birth and affect the epigenome. Our data did not show epigenetic differences in cord blood between cases and controls, but it does not exclude the existence of such differences elsewhere, e.g. in pancreatic cells.

This study was limited to an overall comparison between healthy controls and a heterogeneous group of case children with different first-appearing autoantibodies, seroconversion ages (range 0.5 – 11.6 years), and diagnosis ages (range 1.6 – 18.8 years), who might represent different disease subtypes, the existence of which has been suggested by several studies during the past decade (23). For example, the group of children with IAA as the first-appearing islet autoantibody is characterized by a different HLA-DR-DQ profile and age at seroconversion compared to children with GADA as the first-appearing autoantibody (23,24). The epigenetic profile of newborn infants representing a potential disease subtype, for example children who develop type 1 diabetes at very young age, would be an interesting goal for future studies.

## Data Availability

The datasets generated during the current study will be available in a public repository as soon as the manuscript is accepted for publication in a peer-reviewed journal. The code is available in https://github.com/EssiLaajala/RRBS_workflow, where also the data availability status will be updated.

## ACKNOWLEDGEMENTS

We are grateful to the families for their participation in the DIPP study and the personnel of Turku University Hospital for excellent collaboration, work with the families, and collection of the samples for the study. We also thank the personnel of the HLA laboratory at University of Turku and the personnel of the islet autoantibody laboratory at University of Oulu. We thank emeritus professor Olli Simell for his long-term commitment to the DIPP study. We thank Marjo Hakkarainen and Sarita Heinonen for their excellent technical help. We acknowledge the Turku Bioscience Centre’s core facility, the Finnish Functional Genomics Centre (FFGC) supported by Biocenter Finland, for their assistance. We acknowledge the Finnish Centre for Scientific Computing (CSC) and the computational resources provided by the Aalto Science-IT project.

This study was supported by InFLAMES Flagship Programme of the Academy of Finland (decision number: 337530). R.V. received funding for the DIPP study from the JDRF (grants 1-SRA-2016-342-M-R and 1-SRA-2019-732-M-B), Academy of Finland (grant 308067) and the Finnish Diabetes Foundation. The DIPP Study was also supported by Special Research Funds for University Hospitals in Finland. R.La. received funding from the Academy of Finland (grants 292335, 294337, 319280, 31444, 319280, 329277, 331790), Business Finland and by grants from the JDRF, the Sigrid Jusélius Foundation (SJF), Jane and Aatos Erkko Foundation, Finnish Diabetes Foundation and the Finnish Cancer Foundation. R.La., H.L., M.K., M.O. and J.T. were supported by the Academy of Finland, AoF, Centre of Excellence in Molecular Systems Immunology and Physiology Research (2012-2017) grant 250114 and grant 292482. J.T. was funded by EFSD, Pediatric Research Foundation, and Turku University Hospital Special Governmental Grants. L.L.E. reports grants from the European Research Council ERC (677943), European Union’s Horizon 2020 research and innovation programme (675395), Academy of Finland (296801, 310561, 314443, 329278, 335434 and 335611), and Sigrid Juselius Foundation. Our research is also supported by University of Turku Graduate School (UTUGS), Biocenter Finland, and ELIXIR Finland. E.L. was supported by Turku Doctoral Programme of Molecular Medicine (TuDMM), Finnish Cultural Foundation, and Kyllikki and Uolevi Lehikoinen Foundation.

## Author contributions

E.L. analyzed the data, participated in the interpretation of the results, prepared the figures and tables, and wrote the manuscript. U.U. participated in the interpretation of the results and the drafting of the manuscript. T.G. was responsible for the technical validations by targeted pyrosequencing. O.R. supervised the laboratory experiments and participated in the design of the study setup. V.H., B.R.G., and A.L. participated in the data analysis. M.Ko. and R.K. participated in the technical validations by targeted pyrosequencing. J.M., M.N., and M.V-M. provided the clinical information on the study children compiled by E.L., H.K., and N.L.. H.H., J.I., M.Kn., R.V., and J.T. were responsible for the DIPP study. L.L.E. supervised B.R.G. and A.L.. J.I. provided the samples and was responsible for the DNA isolation and HLA screening of the study children. M.Kn and M.O. initiated and designed the study together with R.La.. R.J.Lu. supervised M.Ko., was responsible for the bisulfite sequencing, and participated in the interpretation of the results. R.V. directed the clinical multicenter DIPP study and was responsible for the DIPP autoantibody laboratory. H.L. supervised E.L. and V.H. for data analysis and participated in the interpretation of the results. J.T. supervised M.V-M. and M.N. and provided the clinical information on the study children. R.La. initiated the study, designed the study setup, supervised the study, participated in the interpretation of the results, and revised the manuscript. All authors contributed to the final version of the manuscript. R.La. is the guarantor of this work and, as such, had full access to all the data in the study and takes responsibility for the integrity of the data and the accuracy of the data analysis.

