## Supplementary Material for "Umbilical Cord Blood DNA Methylation in Children Who Later Develop Type 1 Diabetes"

To a large extent, the content of this supplementary document is also included in a preprint by Laajala et al. (1), which discusses the data analysis workflow in more detail. Most analyses were conducted in R, versions 3.6.1 and 4.0.4 (2). The code is available in https://github.com/EssiLaajala/RRBS_workflow.

### Supplementary methods

##### Sample collection, DNA extraction, and HLA risk class determination

Umbilical cord blood was collected immediately after birth in 3 ml K3-EDTA tubes in the delivery room at the Turku University Hospital and transferred to the DIPP study centre where it was stored at -20°C. After receiving informed consent from the parents, each sample was transferred to the immunogenetics laboratory (destroyed in case informed consent was not received), where it was thawed, and a drop of blood was transferred to a sample collection card for HLA genotype determination. The remaining sample was stored at -20°C. DNA was extracted by applying salting out procedure (3). HLA-DR-DQ genotypes were determined from the DNA on the dried blood spots using assays that were designed to densely probe the genomic regions associated with type 1 diabetes. The genotyping was started from major DQB1 alleles. A hypervariable region on the second exon was further sequenced for individuals with certain DQB1 haplotypes. A detailed description of the genotyping procedure and the risk class definitions have been reported earlier (4,5). The genotypes corresponding to each risk class within this study are:

High (any of the following):
(DR3) - DQA1*05 - DQB1*02 / DRB1*0401 - DQA1*03 - DQB1*0302
(DR3) - DQA1*05 - DQB1*02 / DRB1*0404 - DQA1*03 - DQB1*0302

Moderate (any of the following):
(DR3) - DQA1*05 - DQB1*02 / (DR3) - DQA1*05 - DQB1*02
(DR7) - DQA1*0201 - DQB1*02 / DRB1*0401 - DQA1*03 - DQB1*0302
DRB1*0401 - DQA1*03 - DQB1*0302 / (DR1/10) - DQB1*0501
DRB1*0401 - DQA1*03 - DQB1*0302 / (DR13) - DQB1*0604
DRB1*0401 - DQA1*03 - DQB1*0302 / (DR13) - DQB1*0609
DRB1*0401 - DQA1*03 - DQB1*0302 / (DR16) - DQB1*0502
DRB1*0401 - DQA1*03 - DQB1*0302 / (DR8) - DQB1*04
DRB1*0401 - DQA1*03 - DQB1*0302 / (DR9) - DQA1*03 - DQB1*030
DRB1*0401 - DQA1*03 - DQB1*0302 / DRB1*0404 - DQA1*03 - DQB1*0302

Slightly elevated (any of the following):
(DR3) - DQA1*05 - DQB1*02 / (DR1/10) - DQB1*0501
(DR3) - DQA1*05 - DQB1*02 / (DR13) - DQB1*0604
(DR3) - DQA1*05 - DQB1*02 / (DR8) - DQB1*04
(DR3) - DQA1*05 - DQB1*02 / (DR9) - DQA1*03 - DQB1*0303
(DR7) - DQA1*0201 - DQB1*02 / DRB1*0404 - DQA1*03 - DQB1*0302

DRB1*0401 - DQA1*03 - DQB1*0302 / DRB1*0403 - DQA1*03 - DQB1*0302
DRB1*0404 - DQA1*03 - DQB1*0302 / (DR1/10) - DQB1*0501
DRB1*0404 - DQA1*03 - DQB1*0302 / (DR8) - DQB1*04
DRB1*0404 - DQA1*03 - DQB1*0302 / (DR9) - DQA1*03 - DQB1*0303

Neutral (any of the following):
(DR1/10) - DQB1*0501 / (DR1/10) - DQB1*0501
(DR3) - DQA1*05 - DQB1*02 / DRB1*0403 - DQA1*03 - DQB1*0302
DRB1*0401 - DQA1*03 - DQB1*0302 / (DR14) - DQB1*0503
DRB1*0401 - DQA1*03 - DQB1*0302 / (DR7) - DQA1*0201 - DQB1*0303
DRB1*0404 - DQA1*03 - DQB1*0302 / (DR13) - DQB1*0603
DRB1*0405 - DQA1*03 - DQB1*0302 / (DR7) - DQA1*0201 - DQB1*0303

#### Sample inclusion criteria

Out of 200 umbilical cord blood DNA samples within this analysis, 20 were rejected due to low (< 97 %) bisulfite conversion efficiency, two were excluded due to completely missing hospital metadata, and five were rejected due to inadequate amount or quality of DNA. Out of the 173 remaining DNA samples, 43 are from DIPP case individuals, who have been either diagnosed with type 1 diabetes (N = 34) and/or tested positive for a minimum of two islet autoantibodies on at least two consecutive visits to the DIPP clinic by 2018. The control individuals (N = 79) have remained completely autoantibody-negative throughout the follow-up period (from birth until year 2018, age 15, or their decision to discontinue the study). Samples from individuals with any transient islet antibodies (N = 47) or persistent positivity for only one islet antibody (N = 4) were excluded from the case-control-comparison but included in the analysis of sex-associated differential methylation. The characteristics of the case and control subjects are described in the main text Table 1.

#### Inclusion of maternal data

All available data on the newborn infants were retrieved from Turku University Hospital. One clinical covariate from each group of mutually correlating covariates was included in the differential methylation analysis (described below) to account for potential confounding effects. Pearson correlations greater than 0.3 (in absolute value) with P value < 0.05 or alternatively Fisher’s exact test P value < 0.05 (for pairs of binary covariates) were considered relevant. We prioritized continuous covariates and binary covariates with sufficient sample numbers in both groups. We avoided including covariates that contained missing values or are difficult to measure, such as the duration of delivery phase 1. Further details are in Supplementary Table 1 (below).

#### Reduced representation bisulfite sequencing

DNA concentrations were measured with Thermo Scientific NanoDrop 2000, and the samples were purified with Genomic DNA Clean & Concentrator -10/ ZR-96 Genomic DNA Clean & Concentrator -5 plate kits (Zymo Research, cat. nos D4010 and D4066) according to the protocols for each kit. Library preparation was started from 200 ng of genomic DNA, and *E. coli* genomic DNA (USB, cat. no. 14380) was used as a carrier, 50 ng/library. Library preparation protocol for reduced representation bisulfite sequencing (RRBS) was adapted from (6). To reduce the occurrence of adapter dimers, a lower concentration of adapters (1:10 dilution) was used than recommended by the manufacturer. Bisulfite conversion and sample purification were carried out according to the Invitrogen MethylCode Bisulfite Conversion Kit protocol. Aliquots of converted DNA were amplified by 18 cycles of PCR with Taq/Pfu Turbo Cx Polymerase, a proofreading PCR enzyme that does not stall at uracil. PCR-amplified RRBS libraries were extracted using two subsequent rounds of SPRI bead clean-ups to minimize primer dimers in the final libraries. The high quality of the libraries was confirmed with either Advanced Analytical Fragment Analyzer or Bioanalyzer, depending on the size of the prepared library batch. The concentrations of the libraries were quantified with Qubit® Fluorometric Quantitation, Life Technologies, and only high-quality libraries were sequenced.

The samples were normalized and pooled for the automated cluster preparation which was carried out with Illumina cBot station. The libraries were run in 32 lanes, four-seven samples per lane. The samples were sequenced with Illumina HiSeq 2500 instrument using TruSeq v3 sequencing chemistry. Paired-end sequencing with 2 x 100 bp read length was used with 6 bp index run. The technical quality of the HiSeq 2500 run was high and the cluster amount was as expected. Greater than 85 % of all bases above Q30 was requested. The yields were 18 - 37 million raw paired-end reads per sample. The base calling was performed using Illumina's bcl2fastq2 software, the output of which is of standard fastq format.

##### RRBS data preprocessing and filtering

A more detailed description of the RRBS data analytical workflow can be found in (1). TrimGalore (7) was run with its default parameters for paired-end RRBS data to remove end repair biases, sequencing adapters, and bases with high sequencing error rate (above 1 %). The reads were aligned on GRCh37 (hg19) genome assembly (8,9) and the lambda phage genome simultaneously with Bismark aligner (10). Bismark methylation extractor was run to extract methylated and total read counts for each CpG site in each sample. Bisulfite conversion efficiency was estimated with two methods: 1) based on the total and methylated reads mapped to the completely unmethylated lambda genome and 2) based on cytosines in non-CpG contexts, which are close to 100 % unmethylated in the human genome. Samples with conversion efficiency estimates under 97 % (based on either one of the methods) were excluded from further analysis. After M-bias reports were examined, Bismark methylation extractor was re-run such, that three bases were ignored from the 5’ end of read 2 and one base from the 3’ end of read 2. Finally, the information from both strands was merged for each CpG site.

Single nucleotide polymorphism (SNP) detection was performed by applying BS-SNPer (11) with its default parameters, and the detected SNPs (flagged “PASS”) were removed from the data of each individual (read counts set to NA). To remove most PCR duplication biases, CpG sites with coverage above the $99.9^{th}$ percentile were removed from each sample. CpG sites were completely excluded if they had a low-coverage value (total number of reads < 10) or a missing value (zero coverage, a potential SNP, or extremely high coverage) in at least two thirds of the case subjects or the control subjects.

#### Differential methylation analysis

A generalized linear mixed effects model (GLMM) implemented in R package PQLseq (12) was fit separately for read counts at each CpG site on autosomal chromosomes. A pseudo-count transformation (+1 to methylated and +2 to total read counts) was first applied to all non-missing (total read count > 0) values in the data. The covariates listed in the main text Table 1 were included in the model, based on the criteria specified in Supplementary Table 1. At each CpG site, binary covariates were included only if at least three samples with enough coverage (pseudo-count transformed coverage ≥ 12) were available for each category. Genetic similarities of these 122 unrelated individuals were estimated based on the SNPs detected as described above, and included in the model as random effects via the relatedness matrix, as described by the authors of PQLseq (12). A few missing covariate values (the precise numbers of which are specified in the main text Table 1) were median-imputed, and continuous covariates were Z-transformed. The Wald test *P* values computed within PQLseq were spatially adjusted by a weighted Z-test implemented in the adjust-function of package RADMeth (13) after sorting the CpG sites by chromosome and location. Since the spatially adjusted *P* values were found to be inflated, false discovery rate (FDR) was estimated empirically through a permutation analysis (1). The same workflow was applied to detect differentially methylated CpG sites associated with sex with two modifications: all 173 samples with high-quality RRBS data were utilized (not only the samples from 122 individuals who qualified as cases or controls) and HLA risk was not included as a covariate.

#### Pyrosequencing validation of selected targets

A sex-associated differentially methylated region on the promoter of zona pellucida binding protein 2 (*ZPBP2*) and a class-associated differentially methylated region on an intron of Plakophilin 3 (*PKP3*) were selected for technical validation with targeted pyrosequencing. Four other targets (Supplementary Table 2) had been pyrosequenced before we discovered the P value inflation. For this analysis, 28 case subjects and 30 control subjects were included, based on the following selection criteria: Amount of remaining DNA (determined by Qubit™ dsDNA HS Assay Kit, Invitrogen Ref Q32854), full-term (gestational age ≥ 37 weeks), similar sex distributions in both groups, normal birth weight (2.5 – 4.5 kg), no multiple pregnancies, normal Apgar points (8 – 10), no perinatal asphyxia, vaginal birth and no maternal smoking. PyroMark assay design 2.0 software (Qiagen) was used to design the assays for the methylation validation for the selected targets. Target specific primers designed with the software include primer pair with site-specific biotinylation for target amplification and pyrosequencing primer. The sequencing was done on three to five batches such that a roughly even numbers of male and female case and control subjects were allocated to each batch. The regions of interest were chr17:38024237–chr17:38024290 on hg19 coordinates, including six differentially methylated CpG sites and chr11:400295–chr11:400288, including two CpG sites.

The samples were prepared for pyrosequencing from 200 ng of DNA per sample. Bisulfite treatment was performed using EZ DNA Methylation-GoldTM Kit (Zymo Research cat no D5006) following the manufacturer's instructions. The targets were then amplified using PyroMark PCR Kit (Qiagen cat no 978703). The following PCR conditions were used for amplifications: initial denaturation at 95 °C for 15 min, 45 cycles at 94 °C for 30 s, annealing at 56 °C for 30 s, extension at 72 °C for 30 s and final extension at 72 °C for 10 min. For pyrosequencing reaction, biotin-labeled template strand was used with specific primers. Pyrosequencing was performed with PyroMark Q24 system (Qiagen) with PyroMark Q24® Advanced CpG Reagents (Qiagen cat no 970922). Methylation percentages were extracted using PyroMark Q24 Advanced 3.0.1 software. Supplementary Table 2 summarizes the pyrosequencing and RRBS results for regions that were chosen for pyrosequencing based on Benjamini-Hochberg-corrected spatially adjusted P values before the P value inflation was discovered.

##### Pyrosequencing data analysis

Each methylation proportion was transformed with arcsin(2*proportion-1) before fitting a standard linear model. Besides the sequencing batches, the covariates specified in Supplementary Table 1 were included in the model (excluding covariates that were not applicable, such as maternal smoking which was 0 for all pyrosequenced samples). The model was fit with and without the covariate of interest, and the models were compared with a likelihood ratio test to assess the significance of the covariate of interest at each CpG site. Pyrosequencing data was analyzed using lm and anova, functions in R version 4.0.4. (2).

#### Supplementary results

The results for all pyrosequenced candidate differentially methylated cytosines and regions are presented below. Two CpG sites on the intron of plakophilin 3 (*PKP3*) showed weak evidence of differential methylation between cases and controls. Three empirically obtained null distributions of spatially adjusted P values included some values that were of the same order of magnitude ($10^{-13}$) but not smaller. Targeted pyrosequencing showed that the difference was not significant (Supplementary Figure 1). Supplementary Table 2 summarizes RRBS and pyrosequencing results for genomic regions that would have been considered differentially methylated based on standard criteria (Benjamini-Hochberg-corrected spatially adjusted P value < 0.05) but were declared as false positives based on the empirical FDR control of RRBS results, as well as the pyrosequencing results.

Supplementary Figure 2 shows boxplots for six sex-associated differentially methylated cytosines, three of which reached genome-wide significance in the original RRBS data. This sex-associated region on the promoter of Zona Pellucida Binding Protein 2 (*ZPBP2*) was selected as a positive control to verify that between-group DNA methylation differences could be concordantly quantified by two different technologies.

### Supplementary Figures and Tables

Supplementary Table 1: All variables with available data are within this table. Column 2 indicates whether the variable was included in the differential methylation analysis, which was done with a generalized linear mixed effects model (GLMM) PQLseq.

| **Covariate(s)** | **Included in regression (yes/no)** | **Details and reasons for inclusion/exclusion** |
|---|---|---|
| Age, mother | Yes | The maternal age correlates only with her number of earlier pregnancies and deliveries, and is prioritized over them, since continuous covariates are easier to model than counts. |
| Birth length | No | Correlates with birth weight |
| Birth weight | Yes | Birth weight is included to represent the child's size. It is prioritized over birth height, head circumference and pregnancy duration, since birth weight has the best measurement accuracy. |
| BMI, mother | Yes | This is the pre-pregnancy body mass index. It is prioritized over the body weight, since BMI is more comparable between individuals |
| Class (case/ctrl-status) | Yes | The goal was to study differential methylation associated with this covariate. Case subjects were diagnosed with type 1 diabetes or became persistently positive for at least two islet autoantibodies, whereas control individuals remained completely negative for islet autoantibodies throughout the follow-up. |
| Caesarean section | Yes | The mode of delivery was simplified to vaginal/C-section. C-section only correlates with perinatal asphyxia and is prioritized, since it is simple to define and does not include any measurement uncertainty |
| Age at diagnosis, age at seroconversion, first-appearing autoantibodies | No | Only relevant for case individuals |
| Duration of delivery stage 1 | No | Difficult to measure and includes too many missing values (22 out of 122), for example for all C-section cases. Also correlates with the usage of epidural anesthetic |
| Duration of delivery stage 2 | No | Too many missing values (19 out of 122), for example at all C-section cases. Also correlates with the usage of epidural anesthetic |
| Gestational vaginal bleeding | No | A binary variable, correlates with induced labor |
| Gestational weight gain, mother | Yes | Does not correlate with any other covariate |

| | | |
|---|---|---|
| Glucose tolerance test result, mother | No | A binary variable, correlates with insulin-treated diabetes and includes many missing values (101 out of 122) |
| Head circumference | No | Correlates with birth weight |
| Height, mother | Yes | Included to represent the maternal size together with the BMI. The maternal height correlates only with maternal weight. |
| HLA risk class | Yes | Since the goal was to study differential methylation associated with later disease progression, independent of the well-studied HLA risk, HLA risk was included as a confounding covariate. The original data included four levels (neutral, slightly elevated, moderate, high) but neutral and slightly elevated were merged into one category. In practice, "HLA risk neutral" and "HLA risk high" were modeled as separate binary covariates (moderate was included in the intercept) |
| Induced labor | Yes | Correlates only with gestational vaginal bleeding and is prioritized, since induced labor is simple to define, whereas gestational vaginal bleeding can have different degrees of severity |
| Infant weight when discharged | No | Correlates with birth weight and is not easily comparable between different individuals, since they spend variable amounts of time in the hospital |
| Insulin-treated diabetes, mother | Yes | This can be insulin-treated diabetes of any type (for example gestational diabetes). This covariate is prioritized over neonatal intensive care, neonatal hypoglycemia, and earlier C-section, since insulin-treatment throughout the pregnancy is expected to be more relevant for umbilical cord blood than events that take place before or after the pregnancy. |
| Library preparation batch | Yes | This is a categorical covariate with seven levels (transformed to six binary covariates + intercept). A median number of 16 samples (range 4 – 32) were processed within the same batch. The samples were allocated to the batches, such that each batch contained a comparable group of cases and controls. |
| Low Apgar points | Yes | The 1-minute Apgar points were used here as a binary variable (0=normal, 1=low). Values 7 and lower were considered low |
| Multiple pregnancy | No | This binary variable correlates with birth weight, and the data only includes a few multiple pregnancies. |
| Neonatal hypoglycemia | No | Correlates with maternal insulin-treated diabetes |
| Neonatal intensive care | No | Correlates with maternal insulin-treated diabetes |

| | | |
|---|---|---|
| Number of earlier C-sections | No | The number of earlier Caesarean sections was simplified to a binary variable (0=none, 1=one or more). It correlates with maternal insulin-treated diabetes. |
| Number of earlier deliveries | No | The number of earlier deliveries was simplified to a binary variable (0=none, 1=one or more). It correlates with the maternal age. |
| Number of earlier miscarriages | Yes | The number of earlier miscarriages was simplified to a binary variable (0=none, 1=one or more). There was no reason to exclude this covariate, since it only correlated with the number of earlier pregnancies. |
| Number of earlier pregnancies | No | The number of earlier pregnancies was simplified to a binary variable (0=none, 1=one or more). It correlates with the maternal age. |
| Perinatal asphyxia | No | Correlates with C-section |
| Pregnancy duration | No | Correlates with birth weight |
| Principal components 1 and 2 | Yes | The principal components analysis (PCA) on the coverage-filtered methylation proportion matrix was done after median-imputing remaining missing values for each CpG site. Principal components 1 and 2 were included in the model to represent technical variation. |
| Sex | Yes | Correlates with no other covariate |
| Smoking during pregnancy | Yes | The data included seven mothers smoking throughout the pregnancy and one mother smoking only during the first trimester. This variable was simplified to 0=no smoking, 1=smoking. Covariates related to in-utero conditions were generally prioritized. |
| Transformed month | Yes | This is the month of birth, cosine-transformed ($\cos(2\pi m/12)$, where m is the month as numbers 1 – 12) |
| Umbilical arterial blood pH | No | Too many missing values (18 out of 122) |
| Usage of epidural anesthetic | Yes | This is a binary variable indicating, whether epidural anesthetic was used during delivery stage 1. The value was corrected from 1 to 0 for two individuals with an elective Caesarean section. This variable was prioritized over duration of delivery stages 1 and 2 since usage of epidural anesthetic is simple to define and does not include missing values or measurement uncertainty. |
| Weight, mother | No | Correlates with BMI and height |
| Year of birth (= sample collection year) | Yes | Included to account for possible technical variation related to storage time at -20°C |

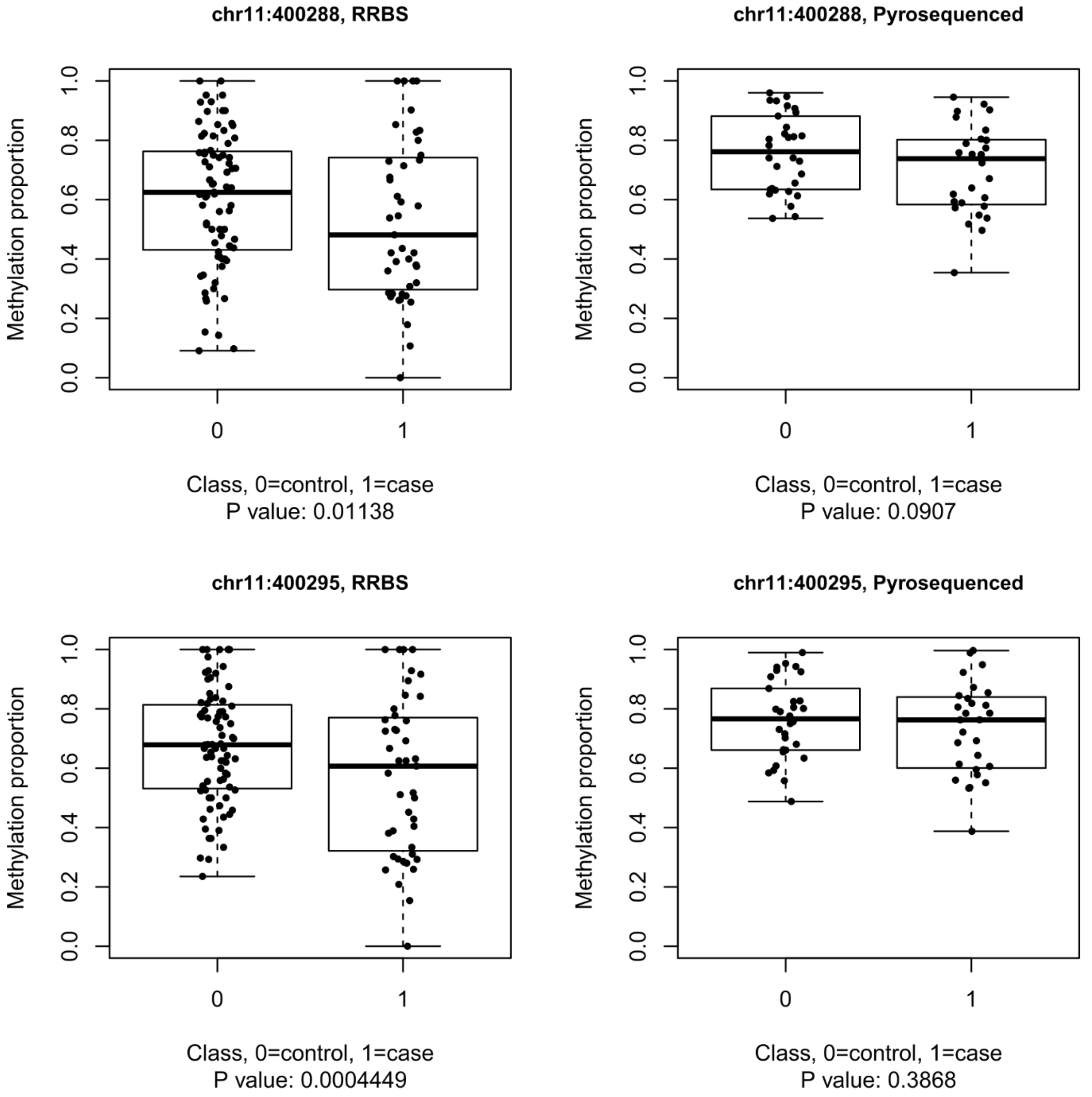


Supplementary Figure 1: Two CpG sites on the intron of Plakophilin 3 (*PKP3*) showed weak evidence of differential methylation between cases and controls (not as individual CpG sites but as part of a region with spatially adjusted P values in the order of $10^{-13}$). However, technical replication by pyrosequencing revealed that the differences were not significant. The RRBS and pyrosequencing results are presented here as boxplots. The raw P values for differential methylation (based on GLMM for RRBS and ordinary linear model for pyrosequencing, separately at each CpG site) are marked below each plot. The midline of each boxplot is drawn at the median, boxes range from the 1st to the 3rd quartile, and whiskers extend to the most extreme values.

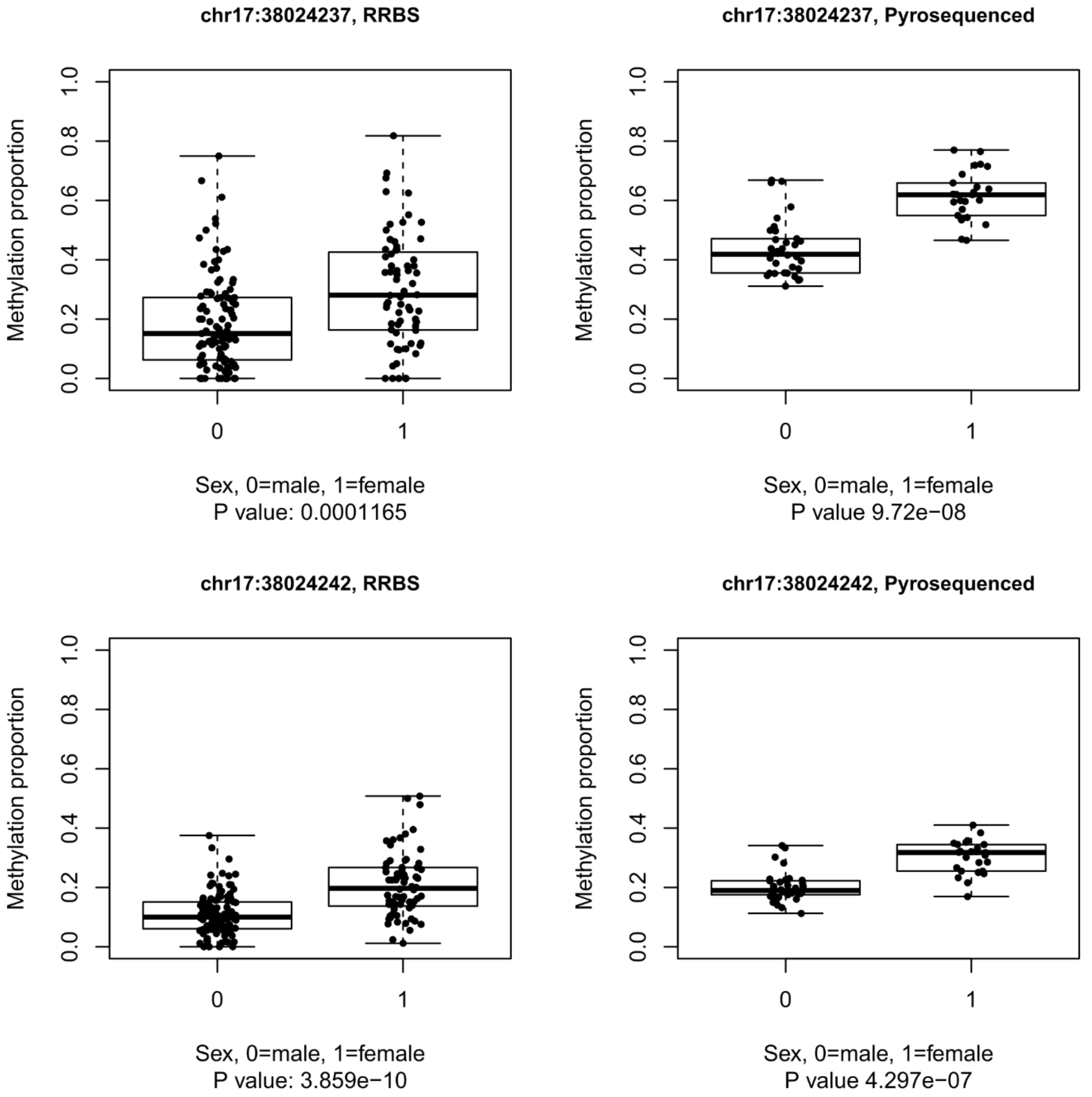
chr17:38024237, RRBS
Methylation proportion
Sex, 0=male, 1=female
P value: 0.0001165
chr17:38024237, Pyrosequenced
Methylation proportion
Sex, 0=male, 1=female
P value 9.72e−08
chr17:38024242, RRBS
Methylation proportion
Sex, 0=male, 1=female
P value: 3.859e−10
chr17:38024242, Pyrosequenced
Methylation proportion
Sex, 0=male, 1=female
P value 4.297e−07

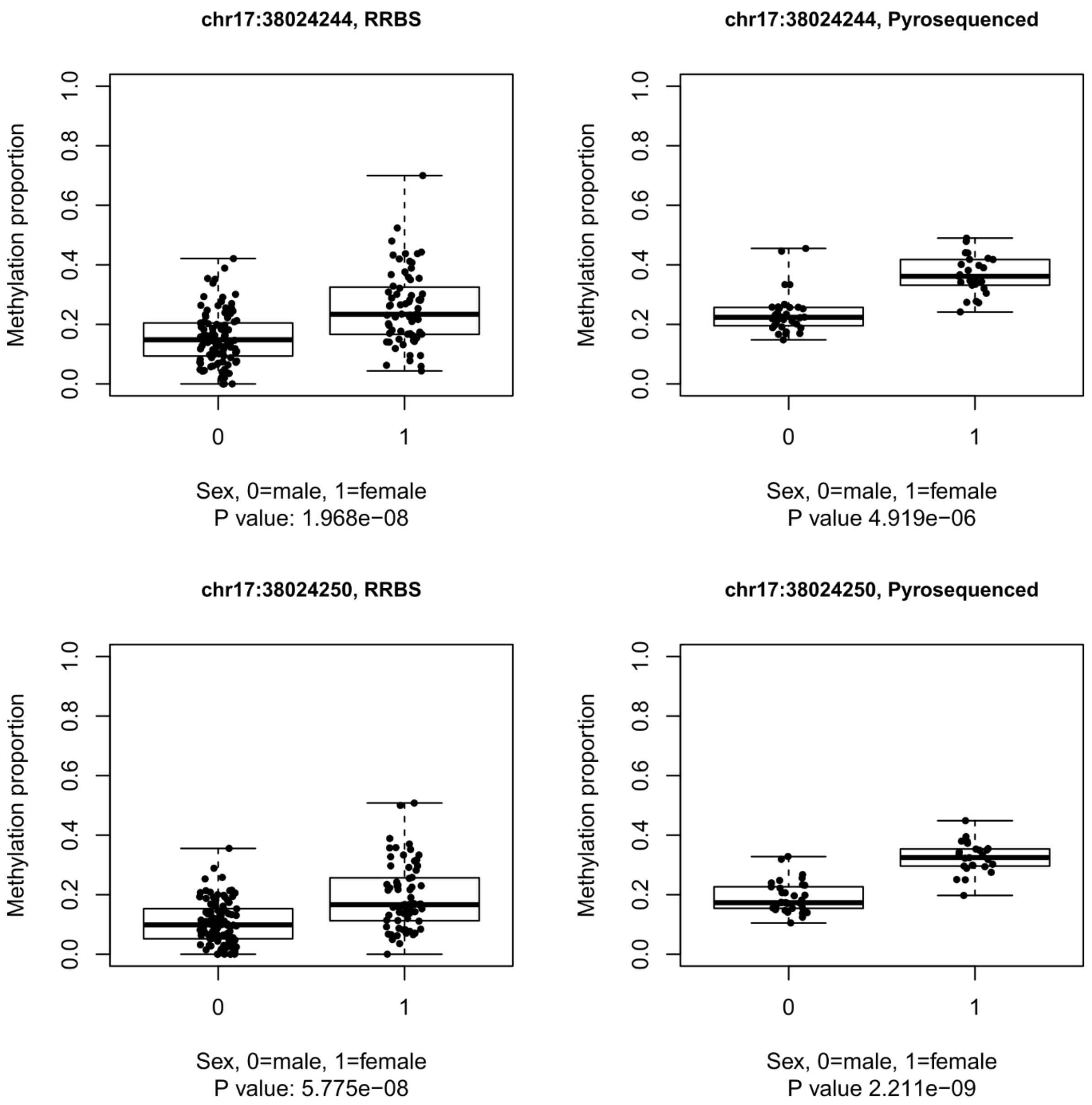
chr17:38024244, RRBS
Methylation proportion
0.0
0.2
0.4
0.6
0.8
1.0
0
1
Sex, 0=male, 1=female
P value: 1.968e−08
chr17:38024244, Pyrosequenced
Methylation proportion
0.0
0.2
0.4
0.6
0.8
1.0
0
1
Sex, 0=male, 1=female
P value 4.919e−06
chr17:38024250, RRBS
Methylation proportion
0.0
0.2
0.4
0.6
0.8
1.0
0
1
Sex, 0=male, 1=female
P value: 5.775e−08
chr17:38024250, Pyrosequenced
Methylation proportion
0.0
0.2
0.4
0.6
0.8
1.0
0
1
Sex, 0=male, 1=female
P value 2.211e−09

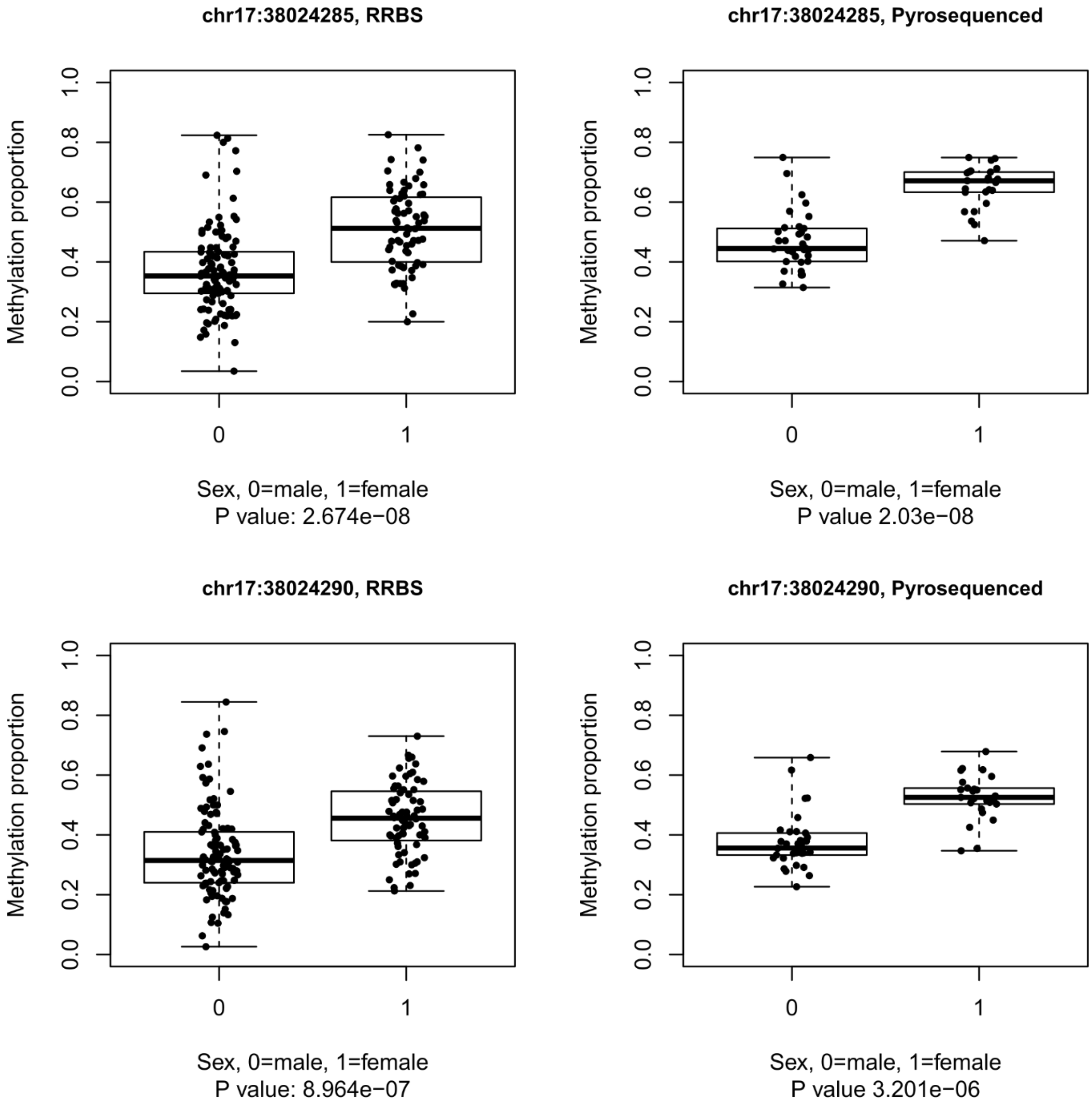


Supplementary Figure 2: Altogether 6 sex-associated differentially methylated cytosines on the promoter of Zona Pellucida Binding Protein 2 (*ZPBP2*) were technically validated by targeted pyrosequencing. The RRBS and pyrosequencing results are presented here as boxplots. The raw P values for differential methylation with respect to sex (based on GLMM for RRBS and ordinary linear model for pyrosequencing, separately at each CpG site) are marked below each plot. The midline of each boxplot is drawn at the median, boxes range from the 1$^{st}$ to the 3$^{rd}$ quartile, and whiskers extend to the most extreme values.

Supplementary Table 2: Summary statistics of genomic regions that were chosen for targeted validation by pyrosequencing based on Benjamini-Hochberg-corrected spatially adjusted P values before the P value inflation was discovered. Based on pyrosequencing, these regions showed no significant differences between the case group and the control group. The empirical FDR control later revealed that the differences were not truly significant in RRBS data either.

| Description of the target(s) | Exon and intron of *GFI1* | Exon of *CHD7* | Intron of *BRSK2* | Intron of *NFATC1* |
|---|---|---|---|---|
| RRBS: candidate differentially methylated region | chr1:92946521-92946810 | chr8:61777859-61777938 | chr11:1412936-1413023 | chr18:77284553-77284613 |
| RRBS: Largest difference between cases and controls | 0.169 | 0.175 | -0.208 | 0.143 |
| RRBS: Smallest raw P value | 0.0074 | 0.0011 | 0.00033 | 0.0047 |
| RRBS: Smallest spatially adjusted P value | 2.90E-07 | 5.00E-06 | 3.70E-05 | 6.40E-06 |
| Pyrosequenced region(s) | chr1:92946620-92946521 and chr1:92946641-92946752 | chr8:61777937-61777909 | chr11:1412927-1413022 | chr18:77284550-77284612 |
| Number of CpG sites within pyrosequenced region(s) | 22 | 4 | 3 | 2 |
| Pyrosequencing: largest difference between cases and controls | 0.052 | 0.023 | -0.097 | 0.028 |
| Pyrosequencing: smallest raw P value | 0.23 | 0.094 | 0.63 | 0.028 |
